# Increase in SARS-CoV-2 RBD-specific IgA and IgG Antibodies in Human Milk from Lactating Women Following the COVID-19 Booster Vaccination

**DOI:** 10.1101/2022.02.23.22271414

**Authors:** Andrea M. Henle

**Affiliations:** Biology Department, Carthage College, 2001 Alford Park Drive, Kenosha, WI, 53140 USA

## Abstract

The CDC recommended a booster dose of the Pfizer-BioNTech Comirnaty (BNT162b2) COVID-19 mRNA vaccine in September 2021 for high-risk individuals. Pregnant and high-risk lactating women were encouraged to receive the booster to obtain potential prolonged protection for themselves and their infants. This study investigated the ability of the booster vaccine to increase IgA and IgG antibodies specific to the receptor binding domain (RBD) of the SARS-CoV-2 spike protein in human milk compared to levels pre-booster. We found a significant increase in both anti-RBD-specific IgA and IgG antibodies in human milk 1-2 weeks after the Pfizer-BioNTech booster and at the study endpoint (60 days post-booster). These results suggest the booster vaccination enhances SARS-CoV-2 specific immunity in human breast milk, which may be protective for infants.

**Key Points:**

- Abs to SARS-COV-2 RBD are detected in blood ≥60 days post-Pfizer-BioNTech booster.
- Human milk Abs to SARS-COV-2 RBD are higher ≥60 days post-booster vs pre-booster.

## Introduction

As of early March 2022, nearly 80 million cases of coronavirus disease of 2019 (COVID-19) were confirmed in the United States and almost 1 million deaths (1). The American Academy of Pediatrics reported that children represented 19% of the cumulative COVID-19 cases and up to 1.5% of those cases resulted in hospitalization (2). While children ages 5 and older are eligible for the Pfizer-BioNTech Comirnaty vaccination, those less than 5 years of age remain ineligible and at risk for infection with severe acute respiratory syndrome coronavirus 2 (SARS-CoV-2), the virus that causes COVID-19. Infants can be protected from diseases such as pertussis and the flu through vaccination of their pregnant or lactating mothers (3, 4). Previous studies have reported elevated levels of SARS-CoV-2-specific IgA and IgG in milk from lactating women following the first and second doses of the Pfizer-BioNtech vaccine (5–9). Therefore, in the absence of FDA-approved COVID-19 vaccinations for infants 6 months or older, maternal vaccination against COVID-19 may offer passive immunity to infants in a similar manner as other vaccines typically administered to pregnant and/or lactating women such as Tdap and Influenza. However, to the best of our knowledge, no one has assessed the antibody response in human milk >2 months beyond the initial two-dose vaccination series for COVID-19.

In September 2021, the CDC recommended a booster dose of the Pfizer-BioNTech Comirnaty (BNT162b2) COVID-19 mRNA vaccine for people ages 65 and older, adults with underlying health conditions, and frontline workers (10). Pregnant and lactating women in these groups were advised to consider the booster for potential prolonged protection for themselves and their infants. We sought to address whether the booster changed the immunoglobulin levels in human milk. This study investigated the ability of the booster vaccine to increase IgA and IgG antibodies specific to the receptor binding domain (RBD) of the SARS-CoV-2 spike protein in human milk compared to the levels pre-booster. We hypothesized that the booster would increase SARS-CoV-2 RBD-specific immunoglobulins in human milk.

## Materials and Methods

### Study design

Participants were recruited through social media posts in academic and regional mom groups. The former groups were targeted because educators and healthcare workers were approved earlier than the general public for the booster and would better align with the timeline of this study. 6 of the 12 participants were recruited prospectively and 6 were recruited approximately one month into the 90-day timeline for the study, but were eligible because they had dated and frozen aliquots of milk from the earlier required time points in the study. Two participants were not able to provide milk samples from 1 day pre-booster (day -1), so their samples from 30-days pre-booster (day -30) were used for the analysis instead. Four participants voluntarily provided milk samples at 90-days post-booster to further extend the analysis timeline. To be eligible for this study, participants needed to be lactating, must have received the standard two-dose Pfizer-BioNTech vaccination series at least 6-months prior, had no known diagnosis or suspected infection for COVID-19, and provide written informed consent. Changes in health status were monitored with a post-study survey. The study was approved by the Institutional Review Board at Carthage College (IRB# 1817816-1).

### Milk and blood sample collection and processing

Participants were instructed to collect ≥2 ounces of milk in the morning at 30-days and 1-day pre-booster, and 7-, 14-, 21-, 30-, 45-, and 60-days post-booster. Samples were immediately stored at −20**°**C until analysis. Upon receipt in the lab, each 2 ounce or greater human milk sample was centrifuged for 25 minutes at 2000 rpm and 4**°**C on a Thermo Scientific Sorvall Legend XTR centrifuge. The aqueous (serum) layer was separated from the fat layer and stored at −20**°**C until the ELISA assay was performed. The negative control was human milk from July 2019.

Participants used an alcohol swab and a finger-prick lancet device to collect up to 200 µl of blood into an BD microtainer in January 2022 (≥60 days post-booster). The blood was allowed to clot for 20 minutes at room temperature and then promptly stored at 4**°**C. Upon receipt in the lab, the blood was centrifuged at 10,000 rpm for 10 minutes at room temperature on an Eppendorf 5424 centrifuge. The serum layer was collected and stored at −20**°**C until the ELISA assay was performed.

### Detection of Abs to SARS-CoV-2 RBD via ELISA

Anti-RBD IgA and IgG levels were assessed via enzyme-linked immunosorbent assay (ELISA) (Ancell Corp.). 96-well plates were coated for 2 hours with 100 µl/well of 4 µg/ml purified RBD-His spike protein (Ancell Corp.). The plates were aspirated and 300 µl/well of blocking buffer (Tris Buffered Saline/Glycine-01% BSA-10% glycerol 0.04% Sodium Azide, pH=7.45) was added for 1 hour at room temperature. Human milk serum was diluted 1:5, and blood serum was diluted 1:100 for IgG analysis and 1:50 for IgA analysis, and incubated on the plates for 1 hour at room temperature with shaking. Plates were washed twice with 300 µl/well Tris Buffered Saline/Glycine, 0.1% BSA, 0.1% Pluronic acid, pH=7.49. Monoclonal mouse anti-human IgG-HRP (ICO-97)(0.8 µg/ml) and mouse anti-human IgA-HRP (Hisa43)(2 µg/ml) antibodies (100 µl/well) (Ancell Corp.) were added to capture the binding signal and the plates were incubated at room temperature with shaking for 1 hour. Plates were washed three times with 300 µl/well TBS. TMB H_2_O_2_ substrate (Ancell Corp.) was used for detection at 450 nm on a Biotek Powerwave X plate reader. All human milk and blood samples were run in triplicate. The negative control for this assay was a human milk sample from July 2019. A positive control and standard curve were generated by serially diluting blood serum obtained from a COVID-19 positive patient in May 2020 into the 2019 negative control human milk sample in duplicate. IgA and IgG levels were converted to units/ml.

### Statistical analysis

Data were analyzed with GraphPad Prism 7. Data were expressed as mean and 95% CIs. A one-way ANOVA with Dunnett’s multiple comparisons test was used to determine differences between the mean levels of IgA and IgG pre-booster (day -1) to all other time points. The significance threshold was p<0.05.

## Results

12 women provided a total of 87 milk samples (Table I). Women were a mean (SD) age of 35.45 (4.17) years and infants 3.58 (1.8) months at the time of booster. The mean (SD) time between the second dose of the vaccine and the booster was 7.01 (0.62) months. 91.67% of women reported a vaccine-related adverse event after the booster, with injection site soreness the most frequent event (75%). 58.33% of women had anti-RBD-specific IgA antibodies and 100% of women had anti-RBD-specific IgG antibodies in their blood at ≥60 days post-booster.

**Table I.** Participant Characteristics.

|  | No. (%) |  |
| --- | --- | --- |
| Study participants, No. | 12 |  |
| Maternal features |  |  |
| White, not Hispanic or Latino or Spanish origin | 12 (100) |  |
| Maternal Age, mean (SD), y | 35.45 (4.17) |  |
| Weight, mean (SD), kg | 83.50 (23.57) |  |
| Height, mean (SD), m | 1.654 (0.075) |  |
| Asthma | 1 (8) |  |
| Antibiotic use in last six months | 2 (17) |  |
| Pregnancy features |  |  |
| Gestational Diabetes | 5 (42) |  |
| Hypertension | 2 (17) |  |
| Gravidity, mean (SD) | 2.75 (1.42) |  |
| Birth week, mean (SD) | 39.30 (0.74) |  |
| First vaccine during pregnancy | 8 (67) |  |
| Pregnancy week at first Pfizer dose, mean (SD) | 22.02 (7.21) |  |
| Pregnancy week at second Pfizer dose, mean (SD) | 25.02 (7.31) |  |
| Infant age when mother received booster, mean (SD), mo | 3.58 (1.8) |  |
| First vaccine after pregnancy | 4 (33) |  |
| Infant age when mother received booster, mean (SD), mo | 10.84 (0.64) |  |
| Time between 2nd vaccine dose and booster, mean (SD), mo | 7.01 (0.62) |  |
| Booster vaccine adverse effects | 11 (92) |  |
| Injection site soreness | 9 (75) |  |
| Injection site rash | 1 (8) |  |
| Injection site swelling | 3 (25) |  |
| Injection site redness | 4 (33) |  |
| Headache | 2 (17) |  |
| Muscle or body aches | 4 (33) |  |
| Joint pain | 3 (25) |  |
| Fatigue or tiredness | 4 (33) |  |
| Fever | 3 (25) |  |
| Chills | 5 (42) |  |
| Syncope | 1 (8) |  |
| IgA positive blood serum at day $\geq 60$ | 7 (58) | |
| IgG positive blood serum at day $\geq 60$ | 12 (100) | |
| Participants represented at each time point, No. (%) | IgA analysis | IgG analysis |
| Day -30 | 8 (67) | 8 (67) |
| Day -1 | 10 (83) | 10 (83) |
| Day 7 | 7 (58) | 9 (75) |
| Day 14 | 10 (83) | 10 (83) |
| Day 21 | 9 (75) | 9 (75) |
| Day 30 | 11 (92) | 11 (92) |
| Day 45 | 11 (92) | 11 (92) |
| Day 60 | 12 (100) | 12 (100) |
| Day 90 | 4 (25) | 4 (25) |

Anti-RBD-specific IgA antibodies in milk significantly increased by day 7; 71.43% of participants had an increased level at day 7 compared to pre-booster (day -1) (Figure 1). IgA levels remained significantly increased at day 60. 41.67% of participants had significantly increased RBD-specific IgA antibodies at day 60. Anti-RBD-specific IgG antibodies in milk significantly increased by day 14; 80% of participants had an increased level at day 14 compared to pre-booster (day -1). The anti-RBD-specific IgG levels remained significantly increased compared to the pre-booster level at 21 days, 45 days, and 60 days. 66.67%, 63.64%, and 75% of participants had significantly increased IgG levels at 21, 45, and 60 days, respectively.

**Figure 1.**
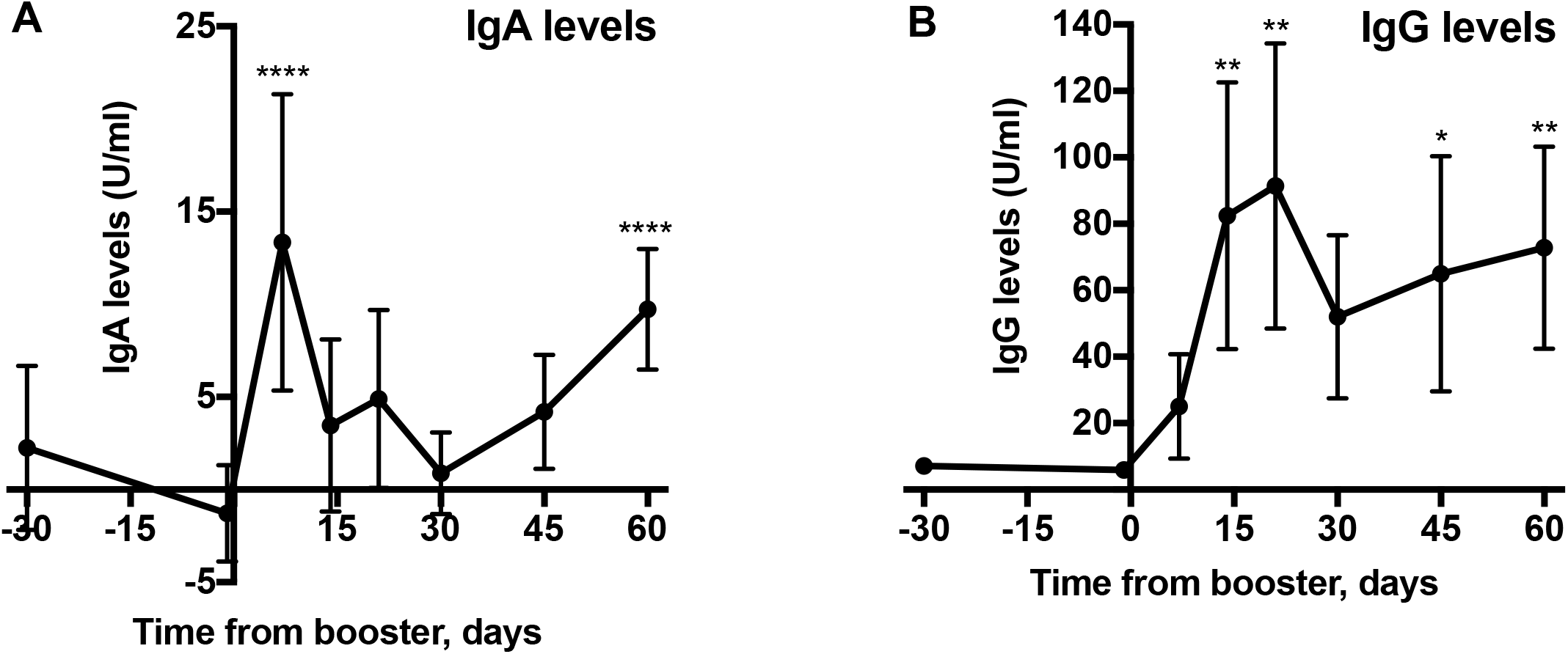
Levels of anti-RBD-specific IgA and IgG in Human Milk Following the Pfizer-BioNTech Booster Vaccine. Each time point is compared to the pre-booster levels of (A) IgA and (B) IgG at day -1. * indicates p<0.05, ** p<0.01, **** p≤0.0001. Each participant’s sample was run in triplicate in one ELISA. Table I indicates the number of participants represented at each time point. Data points represent means; error bars, 95% CIs.

On an individual level, 9/12 participants (75%) had a significant increase in RBD-specific IgA levels at one or more time points post-booster, whereas 12/12 (100%) had a significant increase in RBD-specific IgG at two or more time points post-booster (individual data not shown). Additionally, of the four participants that provided a milk sample at 90-days post-booster, 3/4 (75%) had significantly elevated anti-RBD-specific IgA antibodies (mean 15.24 U/ml) and IgG antibodies (mean 55.67 U/ml) at day 90 compared to pre-booster (day -1) (data not shown).

## Discussion

### Higher levels of IgA and IgG specific to SARS-CoV-2 RBD following the booster

We found a significant increase in anti-RBD-specific IgA and IgG antibodies in human milk following the Pfizer-BioNTech booster. Other studies have reported high IgG levels in human milk following the standard two-dose vaccination against SARS-CoV-2 (5–8), and we observe a similar IgG-dominant effect post-booster. This could be due to the administration method of the vaccination - intramuscular injection may induce a stronger IgG response than other routes which would favor mucosal (IgA) immunity. While IgA is typically considered the predominant immunoglobulin in human milk, previous research suggests IgG in human milk may be important for protection against viral infections such as RSV and HIV (11, 12). Our results, combined with previous studies (5–8), suggest a role for IgG in human milk for immunity to SARS-CoV-2.

One previous study (13) reported that the IgA and IgG antibodies induced following COVID-19 infection are capable of neutralizing SARS-CoV-2 *in vitro*, which suggests that the immunoglobulins induced following the booster vaccination may potentially offer enhanced protection for both the mother and infant.

### Limitations

The sample size in this study was limited and should be expanded to include participants from other races and ethnicities and an age range more representative of the average child-bearing population.

No functional assays were performed to assess the protective activity of the immunoglobulins. Future investigations should use *in vitro* neutralization assays to determine whether SARS-CoV-2 specific antibodies induced post-booster are protective.

This study did not assess immunoglobulins following a booster with the Moderna vaccine or a mix-and-match (Pfizer-BioNTech, Moderna, or Johnson &Johnson) approach to doses 1, 2, and the booster. Future research is needed to assess optimal vaccination strategies (such as the mix-and-match approach) for providing the highest levels of antibodies against SARS-CoV-2 in human milk.

Participants were not tested for SARS-CoV-2 via real-time reverse transcriptase polymerase chain reaction (RT-PCR) as part of this study so it is difficult to determine whether elevated antibody levels could also be attributed to infection (either symptomatic or asymptomatic). However, no participants reported any known exposures to COVID-19 or any positive at-home or RT-PCR test results throughout the duration of the study. Additionally, this study concluded in early January 2022, so most milk sample collection occurred prior to the emergence of the highly transmissible Omicron BA.1 variant of SARS-CoV-2 in the United States. Future studies could quantify the levels of immunoglobulins in human milk that are specific to the SARS-CoV-2 nucleocapsid (N) protein as a way to detect immunity to past infections (since N antigens are not present in the currently available COVID-19 vaccinations).

### Significance

The recent clinical trial which tested the Pfizer-BioNTech vaccination in pregnant women assessed safety and efficacy, but did not directly quantify immunoglobulin levels (14). Healthcare providers and mothers have limited information regarding the ability of the COVID-19 vaccination to induce long-term immunity in human milk. Our research suggests administration of the booster vaccination ≥6 months after the standard two-dose Pfizer-BioNTech vaccination increases SARS-CoV-2 RBD-specific IgA and IgG antibodies in human milk. This may be a source of passive and protective immunity for infants. As of early March 2022, just less than half of the adult population (49.9%) that is booster-eligible in the United States has received their booster vaccine (1). Thus, these results suggest women who are pregnant or lactating and are booster-eligible may wish to consider the booster vaccination at this time for potential protection for their infants.

## Data Availability

All data in the manuscript are available from the corresponding author upon request.

## Acknowledgments

We thank Joan Donner and Paul Everson (Ancell Corp.) for technical assistance with the ELISA assays. We thank Qinzi Ji (Carthage College) for assistance processing the milk samples. We thank Crystal Nelson (Save the Milk) for recommendations and guidance on shipping milk samples. No one was compensated for their contributions.

## Notes

### Competing Interest Statement

The authors have declared no competing interest.

### Funding Statement

This study did not receive any external funding.

### Author Declarations

The Institutional Review Board of Carthage College gave ethical approval for this work (IRB# 1817816-1).

### Summary of Updates

The Introduction section was expanded to provide more background; the Materials and Methods section was expanded and the supplementary methods section was removed; the Results section was expanded and several mistakes were corrected regarding the reported percentages of individuals with increases in IgA levels at day 60 post-booster and IgG at days 14, 21, 45, and 60 post-booster; the Discussion section was expanded; the Acknowledgements section was revised.

